# Immunisation, asymptomatic infection, herd immunity and the new variants of COVID 19

**DOI:** 10.1101/2021.01.16.21249946

**Authors:** Alastair Grant, Paul R Hunter

**Affiliations:** School of Environmental Science, University of East Anglia, Norwich, NR4 7TJ, UK; The Norwich Medical School, University of East Anglia, Norwich, NR4 7TJ, UK

## Abstract

**Objectives:** Is “herd immunity” to COVID-19 a realistic outcome of any immunisation programme with the two main vaccines currently licenced in the UK (Pfizer vaccine BNT162b2 and Astra Zeneca/Oxford vaccine ChAdOx1-S)? More formally, can these vaccines achieve a sufficient level of population immunity to reduce R, the reproduction number of the infection, to below one in the absence of any non-pharmaceutical interventions?

**Design:** The study uses simple mathematical models of the transmission of COVID-19 infection from primary to secondary cases parameterised using data on virus transmission and vaccine efficacy from the literature and the regulatory approval process for the vaccines.

**Results:** In the regulatory approval documents, the efficacy of the Pfizer vaccine is estimated at 0.948 (that for the Moderna vaccine is similar). Efficacy for the Oxford vaccine against primary symptomatic illness is estimated as 0.704, based on pooling of data from two dose regimes. For values of R_0_ similar to those reported during the first months of the pandemic, the simplest analysis implies that reducing the value of R below 1 would require 69% and 93% of the population to be vaccinated with the Pfizer and Oxford vaccine respectively (or achieve a comparable level of immunity through natural infection). However, the new variant of COVID-19 (Lineage B.1.1.7, named Variant of Concern VOC-202012/01) is reported to have an R-value 1.56 (0.92 to 2.28) times higher than the original strain. Vaccinating the entire population with the Oxford vaccine would only reduce the R value to 1.325 while the Pfizer vaccine would require 82% of the population to be vaccinated to control the spread of the new variant.

The Oxford vaccine reduces the incidence of serious illness to a greater extent than it reduces symptomatic illness. But its efficacy against the incidence of asymptomatic infections is lower, reducing its efficacy against *all* infection from 0.704 to 0.525 for the pooled data. Although asymptomatics are less infectious, including them in our calculations still raises R values by 20% or more, from 1.33 to 1.6 for the new variant with 100% vaccination. Neither vaccine is licenced for use in children, and when this is taken into account, this R value rises by a further 37% to 2.2 if the whole adult population is vaccinated. Even the more effective mRNA vaccines may allow the pandemic to persist via transmission amongst children, as current authorisations only allow their use on adults. In the absence of vaccination, R will reduce to 1 when 89% of the population has acquired immunity as a result of previous infection with COVID-19.

**Conclusions:** All currently licensed vaccines provide substantial protection against serious illness to vaccinated individuals themselves. But the Oxford vaccine appears to have relatively low efficacy against asymptomatic infections. Although no comparable data from human trials are available for the mRNA vaccines, non-human primate studies suggest they are better at preventing nasal shedding and so transmission. Herd immunity to COVID-19 will be very difficult to achieve, especially so for the less effective vaccine. The possibility of transmission from vaccinated but infected individuals to vulnerable unvaccinated individuals is of serious concern. There is a strong case for preferring the more effective mRNA vaccines for health and social care workers and those who have contact with large numbers of vulnerable others.

## Introduction

The identification of a new variant of the SARS-CoV-2 virus with higher infectivity indicates even greater threat from the pandemic than previously thought. Appearing as it did at a time of year when coronaviruses are more likely to spread, at least in the northern hemisphere, this adds substantially to the harm that the virus could do to public health and the economic and cultural wellbeing of society. We do, however, have several vaccines being rolled out across different countries. But what if anything does the new variant mean for the effectiveness of the new vaccine? Most commentators do not consider that the new variant will invalidate the effectiveness of the main vaccines (Haynes, Kamath, Lucas, Shon, & Iwasaki, 2021; Xie et al., 2021), but the question of the impact if any this new variant will have on our ability to reach any degree of herd immunity remains. This is needed if immunocompromised people and those who decline immunization are to be protected from the virus.

In this paper we consider whether herd immunity is a realistic outcome of any immunization programme with the vaccines currently licenced in the UK. We also specifically consider how this may change because of the appearance of the new variant and the extent to which the conclusion is altered by the occurrence of asymptomatic infections in vaccinated individuals.

### Parameter estimates

1. R_0_ is the expected reproductive rate of SARS-CoV-2 at the very start of the epidemic, when the whole population was susceptible. A systematic review of estimates of R_0_ from 29 studies included in meta-analysis estimated R_0_ as being 2.87 (95% CI, 2.39–3.44) (Billah, Miah, & Khan, 2020). However, this analysis included several studies that were carried out in January and early February before the more infectious D614G mutation became the more dominant global variant (Korber et al., 2020). Estimates of R_0_ for European countries, generally done later when the D614G mutation had become dominant were often somewhat higher for France (6.32, 95% CI, 5.72–6.99), Germany (6.07, 95% CI, 5.51–6.69), and Italy (3.27 95% CI 3.16–3.38). Two studies for Spain gave (3.56, 95% CI, 1.62–7.82) and (2.26, 95%CI 1.29–3.96). Repeating Billah’s (2020) fixed effects analysis but just for the European studies gives an estimated R_0_ of 3.72 (95% CI = 3.61–3.82).
2. The identification in December of a new variant (Lineage B.1.1.7, and named Variant of Concern VOC-202012/01) suggested an even more infectious variant had arisen (Leung, Shum, Leung, Lam, & Wu, 2020; Public Health England, 2021b; Rambaut et al., 2020; Volz et al., 2020). This new variant is estimated to have a R_0_ that is 1.56 [95%CI: 0.92 - 2.28] times greater than the pre-existing variants.
3. We use the R value of 2.87 for our analysis of the original variant and 2.87 x 1.56 = 4.48 for variant B.1.1.7, but for the reasons discussed in point (1), these are likely to be minimum estimates of the value of R_0_ before and after the emergence of lineage B.1.1.7. If the value of 3.72 from (paragraph 1 above) was used, then R_0_ for the new variant would be 5.80.
4. The efficacy of the Pfizer vaccine BNT162b2 against symptomatic infection is reported to be 94.8% (95%CI 89.8 to 97.6) and that for the Moderna vaccine 94.1% (89.3 to 96.8) (MHRA, 2020b, 2021; Polack et al., 2020). As these values are so similar, we do not explicitly carry out calculations for the Moderna vaccine.
5. The efficacy against symptomatic infection of the Oxford AstraZeneca vaccine (ChAdOx1-S) as stated in the MHRA (2020a) advice to professionals is 70.42% (95% CI 58.84 to 80.63) based on pooling data from two different dose regimes. The efficacy against symptomatic infection in clinical trials of the dose regime of two standard doses that has been approved for use in the UK was 62.1% (41.0 to 75.7%), identified subsequently here as SD/SD. Higher efficacy (90.0%, CI 67.4 to 97.0) was reported for a dose regime in which the first dose was approximately half that intended as a result of changes in methods used to quantify virus concentration in the vaccine (subsequently LD/SD). It is not clear whether these reported differences in efficacy between dose regimes are a consequence of dosage used; the time gap between first and second doses or differences in demographic characteristics of the subjects. Although the effects of the LD/SD regime were analysed post hoc, regulatory approval in the UK has pooled data from both regimes. More details are given in the original peer-reviewed paper and the regulatory approval (MHRA, 2020a; Voysey et al., 2021). The efficacy against asymptomatic infection is discussed below.
6. Quoted efficacies of vaccines are for symptomatic infection and do not exclude the possibility of asymptomatic infection which could still pose a risk of disease transmission. The Oxford AstraZeneca trials included routine swab collection. From this an estimate of the efficacy of the standard dose of the vaccine against asymptomatic infection (and therefore presumably all risk of transmission) was estimated to be just 3·8% (95%CI −72·4 to 46·3) rising to 27.3% (−17.2 to 54.9) for data pooled across both dose regimes (Voysey et al. 2020). Asymptomatics make up 36% of infections in the control group. That the Oxford AstraZeneca vaccine may not prevent asymptomatic infection and therefore stop all transmission was suggested even in early non-human primate studies which found “there was no difference in nasal shedding between vaccinated and control SARS-CoV-2-infected macaques (van Doremalen et al., 2020). We were not able to find similar human data for efficacy of the mRNA vaccines in preventing asymptomatic infection. However, animal studies for both the Pfizer and Moderna vaccines did suggest that these vaccines could stop viral shedding after the 1^st^ day from inoculation and so presumably substantially limit transmission (Corbett, Flynn, et al., 2020; Vogel et al., 2020).
7. In addition, a number of those who received the Oxford vaccine had a positive test for infection with the virus without displaying any of the five most characteristic symptoms of fever ≥ 37.8°C, cough, shortness of breath, anosmia or ageusia. Efficacy stated by Voysey et al (2021) is 36.4% for non-primary symptomatics. If we combine these three groups, the overall efficacy of the SD/SD regime against infection is 39.5% (the figure of 55.7% given by Voysey et al., 2021 for any positive swab includes data for the unlicenced LD/SD regime).
8. Using the figures from Voysey et al, (see table 2), the SD/SD vaccine regime leads to symptomatic cases reducing to 37.9% of that in the control and the combination of asymptomatics and other non-primary is reduced only slightly to 91.3% of the number in the control. The corresponding figures for data pooled across the two testing regimes are 29.6% and 71.3%
9. It is now accepted that asymptomatic but infected individuals can transmit COVID-19 (Buitrago-Garcia et al 2020). But the probability of transmission from someone who is asymptomatic is somewhat less than from someone with symptoms. In the systematic review by Buitrago-Garcia et al (2020) it was estimated that relative risk of transmitting the infection if asymptomatic was 0.35 (95%CI 0.10–1.27) compared to a symptomatic individual. Consequently, any vaccine that reduces the risk of developing symptomatic disease will reduce the transmission even if it does not totally prevent it.
10. There are substantial methodological challenges in characterising the proportion of infections that are truly asymptomatic (Meyerowitz, Richterman, Bogoch, Low, & Cevik, 2020) and estimates vary quite considerably from one study to another. Buitrago-Garcia and colleagues (2020) estimated that overall the proportion of infected individuals who remained asymptomatic throughout their infection was 20% (95%CI 17–25). Voysey et al (2021) have a higher value than this for their control group (36% - see table 2) although this includes some cases where symptom status was not known.
11. None of the three vaccines that have received regulatory approval in the UK are licenced for children. The Oxford and Moderna vaccines are licenced only for those over 18 years old and the Pfizer vaccine for those over 16. Whether vaccination those under these age limits will ever be rolled out is still not clear, although a clinical trial using the Pfizer vaccine with 11-15 year olds has started (Polack et al., 2020). Within the UK, 21% of the population is <18 years old and 19% is < 16 years old (ONS, 2020), so the maximum proportion of the UK population that can currently be offered the Oxford vaccine is 79%, and 81% for the Pfizer vaccine.
12. A recent study amongst health workers (Public Health England, 2021c) found that reinfection of did occur in individuals who were already seropositive or had a previous positive PCR test. Reinfection rates were, however, 83% lower than infection rates in those in which there was no evidence of a previous infection. Only 34.% of reinfected individuals were symptomatic, compared with 78.6% of those infected for the first time.

## Models and results

### Numbers infected

When assessing the efficacy of a vaccine, it is usually assumed that the proportion of individuals who become infective following exposure to the virus is the same as the number who become symptomatic. On this basis, the incidence of both symptomatic and asymptomatic infection amongst those who have been vaccinated with the Pfizer vaccine would be 5% of the rate amongst those who have not been vaccinated. The corresponding value would be 29.6% for the Oxford vaccine, using estimate of efficacy obtained by pooling data across the two dose regimes. However, the efficacy of the Oxford vaccine for asymptomatic infection is lower than that for symptomatic infection. When this is taken into account, the proportion of exposed individuals who become infective rises to 47.5% when asymptomatic infections are included and 60.5% if the efficacy values for the sd/sd regime alone are used (see tables 2 and 3). No data are available on the efficacy of the Pfizer or Moderna vaccines against asymptomatic infection, although the data on the extent to which both of these vaccines stop virus shedding in primate models suggest that it is probably reasonable to assume that their efficacy against asymptomatic infection is similar to that for symptomatic infection (Corbett, Flynn, et al., 2020; Vogel et al., 2020). In the next section, we assess the consequences for disease transmission of the headline efficacy figures and efficacy adjusted for the asymptomatic infections.

**Table 1.** Parameter values. See text for sources. More detail on efficacy is against asymptomatic and non-primary symptomatic infection is provided in tables 2 and 3.

| Parameter | Value | L95%CI | U95%CI |
| --- | --- | --- | --- |
| R0 for old variants, all data | 2.87 | 2.39 | 3.44 |
| R0 for old variants, European data only | 3.72 | 3.61 | 3.82 |
| Ratio between R values for old and new variants | 1.56 | 0.92 | 2.28 |
| Efficacy of Pfizer vaccine for symptomatic infection | 0.948 | 0.898 | 0.976 |
| Efficacy of Oxford vaccine for symptomatic infection, pooled data | 0.704 | 0.588 | 0.806 |
| Efficacy of Oxford vaccine for asymptomatic infection, pooled data | 0.273 | -0.172 | 0.549 |
| Efficacy of Oxford vaccine for symptomatic infection, low dose/standard dose data only | 0.900 | 0.674 | 0.970 |
| Efficacy of Oxford vaccine for asymptomatic infection, low dose/standard dose data only | 0.589 | 0.001 | 0.829 |
| Relative risk of transmission of asymptomatic v symptomatic infection | 0.35 | 0.10 | 1.27 |
| Proportion of unvaccinated infected remaining asymptomatic | 0.20 | 0.17 | 0.25 |
| Proportion of UK population <18 years | 0.21 |  |  |
| Reduction of infection rates in those previously infected | 0.83 |  |  |
| Proportion of reinfected individuals who are symptomatic | 0.343 |  |  |

**Table 2.** Incidence of Primary symptomatic, non-primary symptomatic and asymptomatic infections amongst SD/SD recipients and corresponding control groups from Voysey *et al*. (2021). The final two lines and columns are totals calculated here.

| Group | Vaccinated<br>Incidence rate<br>per 1000<br>person days | Control<br>Incidence rate<br>per 1000<br>person days | Vaccine<br>Efficacy | Vaccinated<br>incidence as<br>proportion of<br>all **control**<br>incidence | Control<br>incidence<br>as<br>proportion<br>of total |
| --- | --- | --- | --- | --- | --- |
| Primary<br>symptomatic -<br>All SD/SD<br>recipients | 56.4 | 148.8 | 62.1% | 0.218 | 0.576 |
| Non-primary<br>symptomatic | 10.3 | 16.3 | 36.8% | 0.040 | 0.063 |
| Asymptomatic<br>or unknown<br>SD/SD<br>recipients | 89.4 | 92.9 | 3.8% | 0.347 | 0.360 |
| Non-primary<br>and<br>Asymptomatic<br>SD/SD<br>combined | 99.7 | 109.2 | 8.7% | 0.386 | 0.423 |
| All categories<br>combined | 156.1 | 258.0 | 39.5% | 0.605 | 1.000 |

**Table 3.** Efficacy calculated by pooling data over both SD/SD and LD/SD dose regimes. Final column is a worst-case scenario for the Pfizer vaccine, assuming efficacy against asymptomatic and non-primary symptomatic infection is the same as that for the pooled data for the Oxford vaccine.

| Group | Vaccinated<br>Incidence rate<br>per 1000<br>person days | Control<br>Incidence<br>rate per 1000<br>person days | Vaccine<br>Efficacy | Vaccinated<br>incidence as<br>proportion of<br>all **control**<br>incidence | Incidence as<br>proportion of<br>all control for<br>Pfizer vaccine<br>efficacy for<br>symptomatics |
| --- | --- | --- | --- | --- | --- |
| Primary<br>symptomatic -<br>All recipients | 44.1 | 149.2 | 70.4% | 0.169 | 0.030 |
| Non-primary<br>symptomatic | 10.3 | 16.3 | 36.8% | 0.039 | 0.039 |
| Asymptomatic<br>or unknown<br>SD/SD<br>recipients | 69.8 | 96.0 | 27.3% | 0.267 | 0.267 |
| Non-primary<br>and<br>Asymptomatic<br>SD/SD<br>combined | 80.1 | 112.3 | 28.7% | 0.306 | 0.306 |
| All categories<br>combined | 124.2 | 261.5 | 52.5% | 0.475 | 0.336 |

### Simple models of disease transmission

In the simplest model of infection transmission:

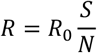

Where the reproductive number R_0_ is the average number of secondary infections per primary case in a population in which all individuals are susceptible; S is the number of susceptibles and N is the total population size. In effect, the number of individuals that are exposed to the virus remains the same but those who are not susceptible, either as a result of vaccination or previous infection develop the disease. For a vaccine that is completely effective, S/N = 1 – v, where v is the proportion of the population that has been vaccinated. If efficacy is less than 100%, then S/N = 1 – E v, where E is the vaccine efficacy, assuming that effective vaccination reduces infectivity to zero and that those in which the vaccine has not been effective are indistinguishable from those who have not been vaccinated (Anderson, Vegvari, Truscott, & Collyer, 2020). The rate of infection will decrease if R < 1. If these assumptions are true, then the proportion of the population that must be vaccinated to control an infection is given by the value of *v* that satisfies:

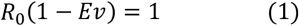

i.e.

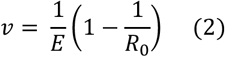

If we use an R_0_ value of 2.87 and a vaccine efficacy of 0.704 (the value on which the regulatory approval was based, MHRA, 2020a), these calculations would indicate that 92.6% of the population need to be vaccinated to reduce the value of R to 1. The headline efficacy of the Pfizer vaccine requires 68.7% of the population to be vaccinated to achieve this. If the new variant increases R_0_ by a factor of 1.56, it would become equal to 4.48. Under these circumstances, the Pfizer vaccine would control the spread of the virus if 81.9% of the population were vaccinated. It is however only authorised for use in those who are 16+, limiting this percentage to a maximum of 81%. If 100% of the population is vaccinated, the Oxford vaccine will only reduce the R value for the new variant to 1.325, rising to 1.98 if only the 79% of the population who are over 18 were vaccinated. So the Oxford vaccine cannot alone control the spread of the virus, even before taking into account that vaccination compliance will be less than complete.

But this analysis does not take into account the lower efficacy of the Oxford vaccine against asymptomatic or non-primary symptomatic infection. The role of disease transmission by asymptomatic cases is a relatively neglected question in epidemiology (Chisholm et al., 2018; Siewe, Greening, & Fefferman, 2020). To give a conservative estimate of the impact of this on transmission, we add the small number of non-primary symptomatics into the asymptomatic category and assume that both have an infectivity that is 0.35 of the value for symptomatics (see above). We assume that overall R_0_ values correctly indicate the reproductive number for the total number of infections, without any bias resulting from under-detection of asymptomatics. The overall R_0_ value can then be decomposed into a component due to asymptomatics and a component due to symptomatics:

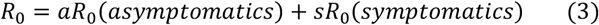

Where *a* and *s* are the proportions amongst those who are infected who are asymptomatic and symptomatic respectively. For an unvaccinated population,

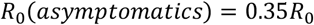

And, using the values of 0.576 and 0.423 for *s* and *a*, based on the characteristics of those infected in the control population of Voysey *et al* (2020), we obtain

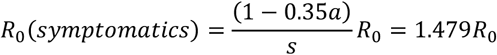

So in an unvaccinated population, transmission by asymptomatics makes up 15% of the total (0.35 x 0.423). Vaccination means that some individuals will not become infected, effectively removing them from the calculation expressed by equation 1. Those who escape infection have an R value of zero, in effect modifying equation (1) so that it becomes:

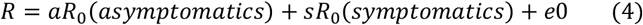

where *e* is the proportion who escape infection and a + s + e = 1. So the consequence of asymptomatic transmission for a range of vaccination scenarios can be assessed by using values of *a* and *r* expressed as proportions of the whole exposed population in equation 3. If vaccine efficacy is identical against symptomatic and asymptomatic infection, then these calculations are equivalent to the simple calculations using equation 1. Results of these calculations for a range of vaccine efficacies and vaccination scenarios are summarised in table 4 and figure 1. More detailed calculations for the most important of these are as follows.

**Table 4.** Summary of R values, proportional reduction in R for vaccinated and percentage of transmission that involves asymptomatic cases for a range of vaccination scenarios, plus natural immunity.

| Scenario | R for new variant with 100% vaccination or infection **(ignoring asymptomatics)** | Factor by which R is multiplied in those vaccinated or infected **(asymptomatics included)** | Percentage transmission that involves asymptomatics | R for old variant (100% vaccination, except top line; **asymptomatics included**) | R for new variant (100% vaccination, except top line; **asymptomatics included**) | R for new variant with all adults vaccinated. **(asymptomatics included)** |
| --- | --- | --- | --- | --- | --- | --- |
| No vaccination |  | - | 15 | 2.87 | 4.4772 | - |
| Oxford, SD/SD | 1.697 | 0.458 | 29 | 1.31 | 2.051 | 2.56 |
| Oxford, efficacy based on pooled dosage data | 1.325 | 0.357 | 30 | 1.02 | 1.598 | 2.2 |
| Oxford, LD/SD regime | 0.448 | 0.125 | 64 | 0.355 | 0.553 | 1.38 |
| Pfizer vaccine efficacy for symptomatics, Oxford vaccine efficacy based on pooled data for asymptomatics | - | 0.151 | 70 | 0.435 | 0.678 | 1.47 |
| Pfizer – assuming same efficacy against asymptomatics as for symptomatics | 0.233 | 0.052 | 15 | 0.149 | 0.233 | 1.124 |
| Immunity after natural infection (based on SIREN study) | 0.386 | 0.125 | 31 | 0.360 | 0.561 | N/A |

**Figure 1.**
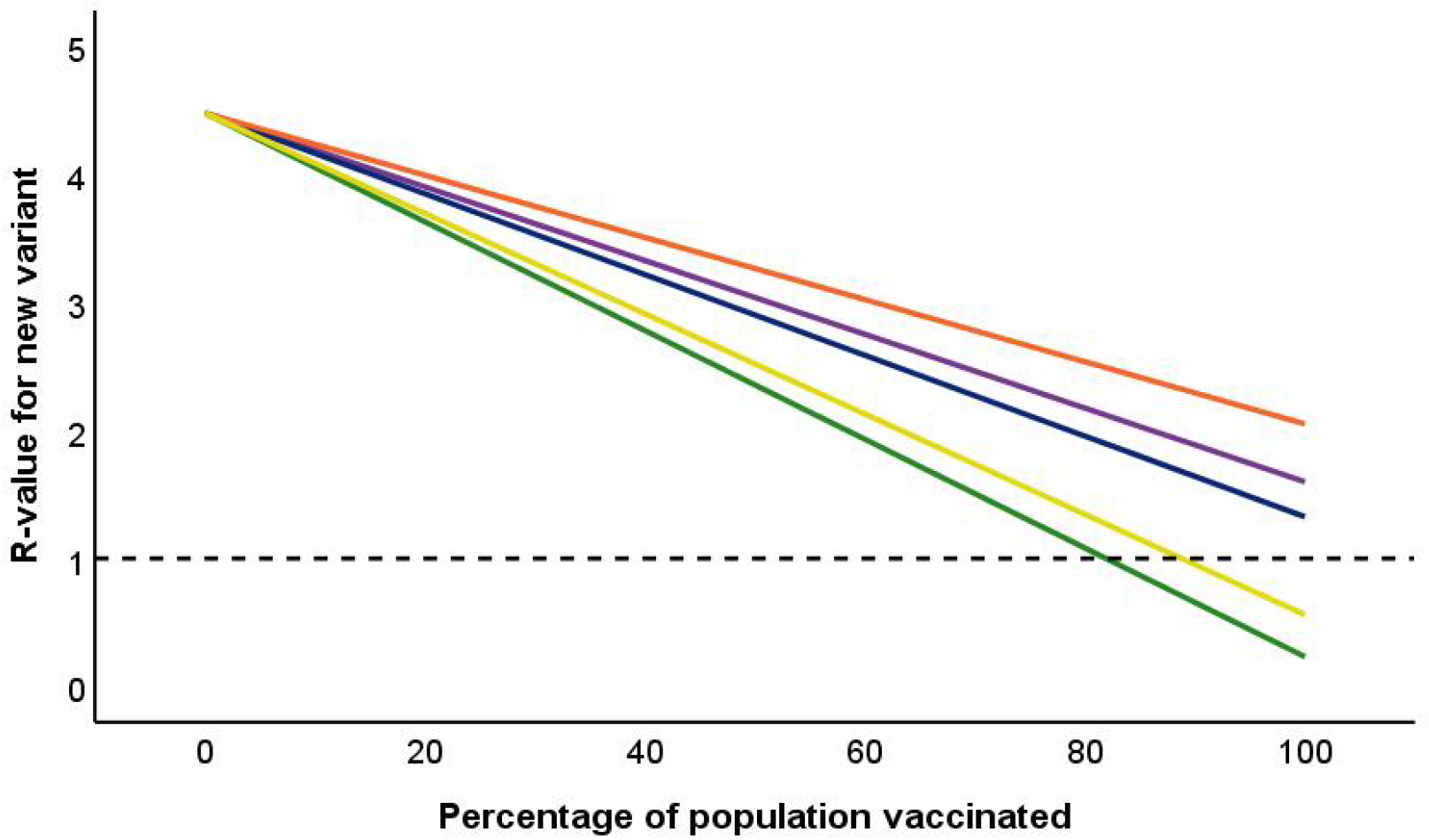
Impact of percentage of population vaccinated on overall R-value for COVID-19. Reference line drawn at R = 1. Green line is Pfizer vaccine; Three upper lines are for Oxford vaccine. Blue: efficacy against symptomatic infection as stated in regulatory approval documents, based on pooling data for SD/SD and LD/SD regime. Purple: same pooled data, but including asymptomatic infection amongst vaccinated individuals. Orange: efficacy for licenced SD/SD regime against both symptomatics and asymptomatics observed in the phase 3 clinical trial (Voysey et al., 2021). Yellow line is equivalent information for immunity in response to natural infection based on data from the SIREN study (Public Health England, 2021c).

Vaccination with the standard dose regime of the Oxford vaccine reduces the number of symptomatics as a proportion of incidence in the control group from 0.576 to 0.218 (0.169 if data from both dosing regimes are pooled). It reduces the proportion of non-primary and asymptomatics from 0.423 to 0.386 (0.306 in the pooled data) and reduces the overall infectivity by a factor of 0.605 (0.475 in pooled data) (see tables 2 and 3). The corresponding values using the efficacy of the Pfizer vaccine for symptomatics would be 0.030, 0.330 and 0.360 respectively.

R for infected vaccinated individuals can be obtained by putting these values for the proportions of symptomatics and asymptomatics amongst vaccinated individuals into equation 1. For the standard dose of the Oxford vaccine, this gives:

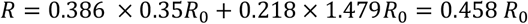

And transmission by asymptomatics makes up 29% of the total. R for unvaccinated individuals remains unchanged. If we denote the proportions of vaccinated and unvaccinated individuals by *v* and *u* respectively, we obtain

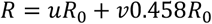

With 100% vaccination, this is equal to 1.31 for the old variant and 2.05 for the new variant, and these figures rise to 1.64 and 2.56 if the 21% of the population under 18 remains unvaccinated. Using the figures for both Oxford vaccine dose regimes combined (on which the regulatory approval was based) gives the R value for vaccinated individuals as:

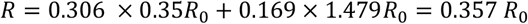

With 100% vaccination, this is equal to 1.02 for the old variant and 1.60 for the new variant and transmission by asymptomatics makes up 30% of the total. With only 79% vaccination, these increase to 1.41 and 2.2 respectively.

If we use the reported efficacies for the Oxford low dose/high dose regime for both symptomatics and asymptomatics, the R value for the new variant with 79% vaccination is reduced to 1.38, and is less than 1 if 100% of the population, including children, were vaccinated. The efficacy of the Pfizer vaccine against symptomatics combined with the efficacy of the Oxford vaccine estimated from both dose regimes for asymptomatics would give an R value equal to 1.47 if all adults were vaccinated. If we assume that the Pfizer vaccine has equal efficacy against asymptomatic and symptomatic infection, then vaccinating all adults would achieve an R-value of 1.12. We can carry out the same analysis using data on the risks of reinfection with the virus amongst those who have previously been infected, and the proportion of asymptomatic individuals amongst these (Public Health England, 2021c). The proportion of transmission by asymptomatic individuals increases to 31% in those who have been infected for a second time. R would have a value of 1 when 89% of the population had been infected (yellow line in Figure 1). If *all* individuals had the level of immunity conferred by previous infection, then R for the new variant would be reduced to 0.561.

## Discussion

These relationships between the percentage of the population that has been vaccinated and R-values are summarised in Figure 1 and table 4. A sufficient level of population immunity could be achievable using the Pfizer (and/or the Moderna) vaccine to reduce R for COVID-19 infections to below 1, and therefore control the epidemic if the whole population were vaccinated. If children remain susceptible, the R-value is predicted to be a little higher than 1. However, moderate levels of immunity in children in response to natural infections in addition to vaccination of adults should be sufficient to reduce R below 1. This should be achievable if immunity in response to previous infection is similar to the 83% reduction observed by the SIREN study (Public Health England, 2021c). By contrast, based on currently reported efficacy the Oxford vaccine allows substantial levels of asymptomatic infection to persist and does not have an efficacy that is high enough to control the more infectious new variant of the virus. This is true even when 100% of the population is vaccinated and the more optimistic efficacy estimates based on pooling data from the two dose regimes are used. Only the efficacy values reported for the low/high dose regime are able to reduce R close to 1, and even then the relatively high incidence of asymptomatic infection will mean that there is a significant risk of those that are vaccinated passing infection on to those that have not been vaccinated or are immunocompromised.

Even very high take up of the Oxford vaccine is unlikely to control the epidemic in the absence of continued non-pharmaceutical interventions to reduce transmission. The Pfizer and Moderna vaccines have efficacy that is high enough to limit the spread of the pandemic, provided a sufficiently high proportion of the population are vaccinated, though it is uncertain that vaccine uptake will be high enough even in people offered vaccine (Freeman et al., 2020). However, the fact that none of the three vaccines is licenced for use on children raises the prospect of continued the pandemic persisting via virus circulation amongst these age groups until a substantial proportion have acquired immunity as a consequence of natural infection.

The aim of the vaccination programme is to “protect those who are at most risk from serious illness or death from COVID-19” (Public Health England, 2021a). Nothing in our analyses would suggest that any of the vaccines currently licenced in the UK would not help substantially to achieve that aim at least in people who have accepted the vaccine. However, the issue remains about how much the current immunisation programme would protect those people who either cannot have the vaccine or elect not to have it. Although vaccination will undoubtedly reduce the risk of symptomatic infection and consequently severe disease, hospitalisation and death, it may do little to protect those people who are unable to have the vaccine or decline the offer. With the available data, herd immunity will be impossible to achieve for the Oxford AstraZeneca vaccine and very difficult to achieve for the mRNA vaccines and consequently unvaccinated vulnerable individuals remain at risk.

This study uses a very simple approach to calculate the consequences of vaccination for rates of disease transmission. At one level this is a weakness as more detailed modelling of infection transmission would give more precise indications of the seriousness of the issues that we highlight. However it is also a strength, as it makes the basis of our argument very clear to see. Our key results follow from the lower efficacy of the Oxford vaccine against symptomatic and its very limited effectiveness against asymptomatic infection, so the overall conclusion would be unlikely to change substantially as a result of the use of more detailed models.

This study includes an explicit consideration of transmission by asymptomatic cases, which has not been considered within the UK vaccine approval process. Evidence from non-human primate studies suggests that this is unlikely to be an important transmission pathway for those receiving the Pfizer or Moderna vaccines, but could be important for the Oxford vaccine as its relative importance increases in vaccinated individuals. Asymptomatic infection raises R-values by more than 20% above those that would be predicted from the efficacy of the Oxford vaccine against symptomatic infection. It could contribute around 30% of transmission using the data on which the UK authorisation for use was based and over 60% of transmission for the dose regime that appeared to have the highest efficacy. This issue has been given relatively little prominence in discussions of the Oxford vaccine, but it reduces efficacy *against infection* below the figures quoted in the regulatory approval documents. Both the blue and purple lines in Figure 1 are based on the pooled dosage data used for regulatory approval, differing only in that the purple line includes transmission by asymptomatics. This also has important implications for the potential for transmission by those who have large numbers of contacts or health and social care staff as it reduces the probability that an individual realises that they are infectious. There is, therefore, a strong case that health and social care staff should be given a vaccine that has high efficacy against both asymptomatic and symptomatic infection. A vaccine with low efficacy against asymptomatic infections would be of less value in reducing the incidence of nosocomial infections.

The Oxford vaccine appears to give a good level of protection against severe disease, as indicated by data on hospital admissions and deaths amongst those who have been vaccinated (Voysey et al., 2021). So there are substantial benefits to the vaccinated individual themselves. It may be much less effective at restricting disease transmission, so even quite high population levels of vaccination offer only limited protection to individuals who have not been vaccinated or have a low immune response to vaccination in the absence of additional non-pharmaceutical interventions. In the medium term, the higher levels of infection that will still be able to circulate in a population that has been vaccinated largely using the Oxford vaccine will provide a much greater opportunity for the virus to evolve increased transmission efficiency or to escape control by the immune response induced by the vaccine (c.f. Antia, Regoes, Koella, & Bergstrom, 2003; Eguia et al., 2020; Nelson et al., 2021). So re-vaccinating using a vaccine with higher efficacy will be a very high priority.

### Unanswered questions and future research include

It would be helpful to have a more detailed understanding of the extent to which the Pfizer and Moderna vaccinations limit the occurrence of asymptomatic infections. To date, the evidence that is available is only circumstantial (although some information on this will come from post-authorisation monitoring-Public Health England, 2020).

More work is needed to understand whether the relatively low efficacy of the Oxford vaccine, particularly against asymptomatic infection, is a consequence of the use of an adenovirus vector; the precise details of the antigen used or some other feature of the vaccine. The Pfizer vaccine has achieved efficacy of approximately 95%, even though both this and the Oxford vaccine use the full length spike protein as the antigen. However, the former has introduced two proline substitutions to lock it in prefusion conformation (Polack et al., 2020) and the Moderna vaccine also uses a similar modified spike protein (Corbett, Edwards, et al., 2020). By contrast, the Oxford vaccine “expresses a codon-optimised coding sequence for the spike protein” (Folegatti et al., 2020; van Doremalen et al., 2020) which, presumably, undergoes trimerisation. Another possible explanation is that efficacy is reduced by an immune response to the adenovirus vector when the second dose is given, something which might be reduced by using two different adenovirus vectors for the two vaccine doses, as in the Sputnik V vaccine (Rawat, Kumari, & Saha, 2021). More detailed work is also needed on the effect of dose regime on the efficacy of the Oxford vaccine including the extent to which the apparent higher effectiveness of the LD/SD regime was due to the dosages themselves or to the time interval between doses. There is some evidence that an increased time interval between doses increased both immune response after the second dose and efficacy, so alterations in the vaccination regime may provide a route to increase its efficacy closer to the value reported for the LD/SD regime in the phase 3 clinical trial (MHRA, 2020a; Voysey et al., 2021) which would greatly increase the extent to which it could control the spread of infection.

As discussed, our analyses suggest that vaccination will not be able to guarantee protection of those people who either cannot have vaccine or decline the offer. If herd immunity is not achieved, they will remain at risk of infection, severe illness and death. How we as a nation deal with this remaining group of susceptible vulnerable individuals is not clear. Will we need to maintain some of the restrictions currently in place to continue to protect these people?

## Data Availability

Data are taken from published papers and sources. All these are cited in the manuscript.

